# Two mosquito salivary antigens demonstrate promise as biomarkers of recent exposure to *P. falciparum* infected mosquito bites

**DOI:** 10.1101/2024.04.20.24305430

**Authors:** Sarah Lapidus, Morgan M. Goheen, Mouhamad Sy, Awa B. Deme, Ibrahima Mbaye Ndiaye, Younous Diedhiou, Amadou Moctar Mbaye, Kelly A. Hagadorn, Seynabou Diouf Sene, Mariama Nicole Pouye, Laty Gaye Thiam, Aboubacar Ba, Noemi Guerra, Alassane Mbengue, Hamidah Raduwan, Inés Vigan-Womas, Sunil Parikh, Albert I. Ko, Daouda Ndiaye, Erol Fikrig, Yu-Min Chuang, Amy K. Bei

## Abstract

**Background:** Measuring malaria transmission intensity using the traditional entomological inoculation rate is difficult. Antibody responses to mosquito salivary proteins such as SG6 have previously been used as biomarkers of exposure to *Anopheles* mosquito bites. Here, we investigate four mosquito salivary proteins as potential biomarkers of human exposure to mosquitoes infected with *P. falciparum*: mosGILT, SAMSP1, AgSAP, and AgTRIO.

**Methods:** We tested population-level human immune responses in longitudinal and cross-sectional plasma samples from individuals with known *P. falciparum* infection from low and moderate transmission areas in Senegal using a multiplexed magnetic bead-based assay.

**Results:** AgSAP and AgTRIO were the best indicators of recent exposure to infected mosquitoes. Antibody responses to AgSAP, in a moderate endemic area, and to AgTRIO in both low and moderate endemic areas, were significantly higher than responses in a healthy non-endemic control cohort (p-values = 0.0245, 0.0064, and <0.0001 respectively). No antibody responses significantly differed between the low and moderate transmission area, or between equivalent groups during and outside the malaria transmission seasons. For AgSAP and AgTRIO, reactivity peaked 2-4 weeks after clinical *P. falciparum* infection and declined 3 months after infection.

**Discussion:** Reactivity to both AgSAP and AgTRIO peaked after infection and did not differ seasonally nor between areas of low and moderate transmission, suggesting reactivity is likely reflective of exposure to infectious mosquitos or recent biting rather than general mosquito exposure. Kinetics suggest reactivity is relatively short-lived. AgSAP and AgTRIO are promising candidates to incorporate into multiplexed assays for serosurveillance of population-level changes in *P. falciparum*-infected mosquito exposure.

## Background

Despite control and elimination efforts, the global malaria burden remains high. *Plasmodium falciparum*, transmitted by *Anopheles spp*. mosquitos, causes the most severe manifestations of disease[1]. Accurate transmission measures are important to estimate disease burden and observe changes in transmission risk. Common metrics of malaria transmission include human biting rate (HBR, the rate humans are bitten by mosquitoes) and sporozoite rate (the proportion of mosquitoes with sporozoites in their salivary glands), the product of which is the entomological inoculation rate (EIR), a measure of the number of infectious mosquito bites a person receives over a period of time[2]. Though EIR is considered the gold standard malaria transmission metric, EIR is costly and difficult to routinely and systematically estimate (especially in low transmission areas), may be imprecise, and may differ based on mosquito catch methods[2].

Serological approaches assessing human antibodies to *Plasmodium* antigens have been used previously as biomarkers for malaria burden and changes in transmission, which has been especially useful in low transmission settings[2-7]. Antibodies to mosquito salivary gland extracts (SGE) have also been investigated as biomarkers of exposure to mosquito bites. Studies have shown IgG responses to SGE correlate with mosquito density[8] and host malaria infection status[9, 10].

Beyond whole SGE, specific mosquito salivary antigens (MSAs) have used as serological biomarkers for *Anopheles* HBR and malaria transmission[11]. Individual proteins investigated as potential biomarkers include SG6[7, 12-18] and the D7 protein family[19]. Antibodies to SG6 in particular have correlated with human exposure to malaria vector bites[18], including marking the heterogeneity of malaria exposure in different geographical settings and between rainy and dry seasons[7, 12], and marking changes of malaria transmission in response to malaria control interventions[13].

This study investigates four additional MSAs with potential to serve as biomarkers specifically for infected mosquito exposure, based on their reported characteristics in pathogenesis studies in human and/or mice *Plasmodium* transmission studies: *Anopheles gambiae* Sporozoite Associated Protein (AgSAP, AGAP004803), *A. gambiae* TRIO (AgTRIO, AGAP001374), mosquito gamma-interferon–inducible lysosomal thiol reductase (mosGILT, AGAP004551), and Sporozoite Associated Mosquito Saliva Protein 1 (SAMSP1, AGAP013726)[20]. Three of these proteins, AgSAP, mosGILT, and SAMSP1, were found to be directly associated with *Plasmodium* sporozoites during transmission using mass spectrometry[20].

The first protein, AgSAP, is expressed in *A. gambiae* salivary glands and the midgut, and has increased expression in the salivary glands of mosquitoes infected with *P. berghei*[21]. AgSAP binds to both *P. falciparum* and *P. berghei* sporozoites, and though AgSAP does not affect sporozoite viability directly, AgSAP knockdown mosquitoes transmitted *P. berghei* less efficiently to mice whereas sporozoites incubated with AgSAP had higher *P. berghei* liver burden[21]. IgG antibodies to AgSAP are elicited by mosquito bites in mice, and additionally, mice immunized with AgSAP and exposed to infectious mosquito bites had a lower liver parasite burden[21]. In humans, people in a malaria-endemic area in Senegal had higher IgG reactivity to AgSAP than people in a non-endemic area[21].

The second protein, mosGILT, binds to the *P. berghei* and *P. falciparum* sporozoite surface, and *P. berghei* and *P. falciparum* sporozoites incubated with recombinant GILT traversed fewer hepatic cells and dermal fibroblasts *in vitro*[20]. Additionally, *P. berghei* sporozoites incubated with recombinant GILT led to a lower murine liver parasite burden[20]. Furthermore, mice immunized with recombinant GILT and challenged with *P. berghei-* infected mosquitoes had higher liver parasite burdens[20].

For the third protein, SAMSP1, recombinant SAMSP1 proteins bind to *P. berghei* sporozoites and enhance sporozoite gliding and hepatocyte cell traversal *in vitro*[22]. Mice receiving SAMSP1 antiserum or IgG purified from SAMSP1 antiserum had a significantly lower *P. berghei* parasite liver burden after *P. berghei* sporozoites were administered via intradermal inoculation[22]. Moreover, active immunization with SAMSP1 lowered the liver burden of mice exposed to *P. berghei*–infected mosquitoes[22]. In humans, people in a malaria-endemic area in Senegal had higher IgG reactivity to SAMSP1 than people in a non-endemic area[22].

The fourth protein, AgTRIO, was identified from IgG purified from SGE antiserum, which was used to probe a cDNA yeast surface display library to identify genes encoding *A. gambiae* proteins with putative signal sequences that could be secreted into saliva[23]. AgTRIO was confirmed to be secreted into saliva, and furthermore, female *P. berghei*-infected *A. gambiae* salivary glands had higher AgTRIO protein levels than uninfected mosquito salivary glands[23]. Additionally, mice immunized with AgTRIO had reduced burden of *P. berghei* in their livers and lower parasitemia when infected by *A. gambiae* mosquito bite[23]. Sporozoites from mosquitoes with RNA interference-mediated AgTRIO silencing colonized mouse livers less effectively[24]. AgTRIO has been investigated as a possible vaccine candidate, with evidence showing mice injected with monoclonal AgTRIO antibodies[25] or immunized with an AgTRIO mRNA-lipid nanoparticle[26] were protected against *P. berghei* infection. Interestingly, people from a malaria endemic area in Senegal did not have significantly higher IgG responses to AgTRIO than people in a non-endemic area[23].

In summary, AgSAP, AgTRIO, and SAMSP1 are associated with increasing *Plasmodium* infection (or alternatively, immunization with these proteins is associated with decreasing infection). Conversely, mosGILT decreases *Plasmodium* infection, and active immunization with mosGILT increases infection. Since these proteins were identified from pathogenesis studies, they were highlighted because of their physical association with *Plasmodium* sporozoites, and thus can be investigated as potential biomarkers *Plasmodium-*infected mosquito exposure regardless of the direction of their effect on *Plasmodium* infection. Importantly, AgSAP and AgTRIO show increased expression in *Plasmodium-*infected mosquitos, leading to our hypothesis that serological responses to these proteins may be higher after exposure to infected versus uninfected mosquitoes.

Here, we leverage a multiplex assay to investigate the human humoral IgG response to sporozoite-associated MSAs as quantitative biomarkers for *Plasmodium*-infected mosquito bite exposure. Multiplex approaches are highly sensitive, allow for detection of multiple serological markers using small sample volumes[27], and are useful for high-throughput serological analyses for disease surveillance[4, 28, 29]. We investigate serological responses to these four MSAs in longitudinal and cross-sectional cohorts from areas of different malaria endemicity, and among people with and without malaria infection. In seeking to validate alternative approaches to standardize and facilitate measure of malaria exposure, here we sought to determine whether cohorts from areas of higher malaria endemicity, and people with active or recent infection, have higher serological responses to these antigens.

## Methods

### Recombinant protein expression

Expression of recombinant mosGILT and SAMSP1 has been described previously[20, 22]. Briefly, recombinant mosGILT and SAMSP1 were expressed using a *Drosophila* expression system based on the expression vector pMT/BiP/V5-His (Invitrogen, CA) and then transfected into *Drosophila* S2 cells with pCoHygro using the Calcium Phosphate Transfection Kit (Invitrogen, CA). Stable cell lines were selected by adding 300 μg/ml hygromycin-B and protein expression was induced by adding copper sulfate to the medium to a final concentration of 500 μM.

The AgTRIO and AgSAP sequences were designed for optimal expression using baculovirus and then subcloned into pFastBac1[24]. Each plasmid was transfected into Sf9 cells with the transfection reagent Cellfectin II (Thermo Fisher Scientific), and then the baculovirus was collected. The Sf9 cells were infected with baculovirus to express AgTRIO or AgSAP, and the protein expression was confirmed by Western blotting. The protein was purified from the cell lysate using a Ni-NTA resin column and then filtered through a 0.22-μm-pore-size filter. The purity was confirmed by SDS-PAGE gel (Figure S1) and Western blot (data not shown) analyses. The protein concentration was determined using a BCA protein assay with BSA as the standard (Thermo Fisher). All expression and purification experiments were performed by GenScript USA, Inc.**Coupling antigens to beads:** MagPlex COOH-microspheres (Luminex Corp., Austin TX) were conjugated to each of the purified antigens, using standard coupling conditions outlined in the xMAP cookbook. Optimal coupling was determined by serial dilution of each protein using the anti-histidine-Biotin antibody (Abcam, ab27025) as all purified contain c-terminal his-tags. Optimal protein concentration for coupling was determined to be 0.3ug of each protein for 1ml of beads. AgSAP was coupled to microsphere region 51, AgTRIO to region 52, SAMSP1 to region 53, and mosGILT to region 54. Bead regions were intentionally chosen to allow multiplex with other existing platforms[4, 30, 31]. Conjugated proteins were validated using sera from immunized and mosquito-exposed mice[20, 22, 23].

### Multiplex assay

We tested IgG responses to AgSAP, AgTRIO, mosGILT, and SAMSP1 in a magnetic bead-based assay on the Bio-rad Multiplex using the CDC multiplex assay procedure[32]. Plasma samples and controls were diluted 1:400 in assay buffer (PBS, 0.05% Tween20, 0.5% BSA, 0.5% PVA, 0.5% PVP, 0.02% NaN_3_, 5% casein). For DBS samples, 6 mm punches were eluted in 200 μl of assay buffer[4, 32]. Since this results in the equivalent of a 1:40 plasma dilution, the eluted DBS were further diluted 1:10 in assay buffer for a final equivalent concentration of 1:400.

Five hundred beads per bead region for each of the four antigens in 50 μl assay buffer were added to each well, the plate was washed twice in PBS with 0.05% Tween20, and 50 μl of diluted sample (plasma or DBS) was added. 50 μl of 0.4% biotin labeled anti-human IgG (Southern Biotech), 0.16% biotin labeled anti-human IgG4 (Invitrogen), and 1% Streptavidin-PE (Invitrogen) in assay buffer was added to each well. Plates were agitated 60 minutes at 700 rpm, washed 4 times, and 50 μl of assay buffer was added per well to ensure unbound proteins were eliminated. Plates were further agitated 30 minutes at 700 rpm, washed twice, and 100 μl of PBS was added to each well. Plates were agitated at 1000 rpm for 30 seconds and then then read on the Bio-Plex 200 in combination with Bio-Plex Manager software version 6.1 (Bio-Rad Laboratories).

Samples were tested in duplicate and the average mean florescent intensity (MFI) was calculated by: MFI_ave_ = (MFI_1_ + MFI_2_ - MFI_blank1_ - MFI_blank2_)/2, where MFI_1_ and MFI_2_ represent the sample duplicates and MFI_blank1_ and MFI_blank2_ represent the blank duplicates. Positive controls (pooled plasma from Mali and pooled plasma from Kédougou, Senegal) and negative controls (healthy, unexposed US controls) were included on each plate.

#### ELISA

On a subset of samples, a standard enzyme-linked immunosorbent assay (ELISA) with AgSAP and AgTRIO IgG was conducted as described previously for comparison with the multiplex cytometric bead assay[22, 23]. Plates were read at 450nm and 570nm, and results of each well was calculated as the result at 450nm minus the result at 570 to remove background. Average optical density (OD) was calculated by: OD_ave_ = (OD_1_ + OD_2_ - OD_blank1_ - OD_blank2_)/2, where OD_1_ and OD_2_ represent the sample duplicates and OD_blank1_ and OD_blank2_ represent the blank duplicates. Plates were tested with positive controls (pooled plasma from Mali and pooled plasma from Kédougou, Senegal) and negative controls (healthy US controls).

### Statistical Analysis

To compare reactivity of different groups and reactivity at different timepoints for longitudinal data, geometric means and geometric mean 95% confidence intervals are reported. For statistical analysis, MFI values were log transformed and analyzed using a linear mixed effects model using Restricted Maximum Likelihood accounting for repeated subjects. Longitudinal results from Thiès and from Kédougou were tested separately. Models showing at least one group differed significantly were then compared with a post-hoc test using estimated marginal means and adjusting p-values for multiple comparisons using Tukey’s method. An alpha of 0.05 was used.

To compare the multiplex results of DBS and plasma samples, Pearson correlations between log transformed MFI values from plasma and DBS sampled collected from the same individuals at the same timepoints were calculated.

To compare results tested by ELISA and the multiplex platform, Spearman correlations of samples tested by both platforms were performed for AgSAP and AgTRIO. Spearman correlations were also performed comparing multiplex results for AgSAP and AgTRIO antigens.

For AgSAP, cutoffs for positivity were calculated using values from the negative HCW cohort. Values were log transformed for normality, and the mean + 2 standard deviations was calculated and then exponentiated to determine a cutoff for positivity. For AgTRIO, cutoffs for positivity were determined from a fixed mixture model (FMM) from a bimodal distribution[33]. Pairwise comparisons for the proportions of positive samples between different groups was compared using Fisher’s exact test, with pairwise comparisons chosen a priori and avoiding comparisons between groups with the same individuals to avoid repeated measures. P-values were adjusted with a Benjamini-Hochberg procedure to correct the false discovery rate. Analysis was conducted and figures were made in R 4.2.2 and GraphPad Prism 9.3.0

## Results

### Demographics

A total of 164 samples were tested from Senegal, with 33 from the low transmission area of Thiès (with an EIR <5[34]), all infected with malaria at the time of enrollment, and 131 from the moderate transmission area of Kédougou (with an EIR ∼250[35]), 72 infected and 59 uninfected at the time of enrollment. The Thiès cohort was followed for 2 years and includes an average of 6.8 samples per individual. Samples from Kédougou were all collected during the malaria transmission season. Uninfected samples came from a cross-sectional study (Table 1).

**Table 1.** Demographics of cohorts.

| Cohort |  | Thiès<br>Infected<br>(Longitudinal) | Kédougou Infected<br>(Cross-sectional and<br>longitudinal)<sup>a</sup> | Kédougou<br>Transmission<br>(Cross-sectional) |
| --- | --- | --- | --- | --- |
| **Malaria transmission**<br>**intensity of region** |  |  |  |  |
| EIR (infectious bites per<br>person per year) |  | <5[34] | ~250[35] | ~250[35] |
| **Demographics** |  |  |  |  |
| Age in years (Mean [Range]) |  | 10.6 (5, 15) | 20.4 (2, 74) | 21.3 (1, 74) |
| Sex | Female (N [%]) | 0 (0%) | 32 (44.4%) | 30 (50.8%) |
|  | Male (N [%]) | 33 (100%) | 38 (52.8%) | 29 (49.2%) |
|  | Unknown (N [%]) | 0 (0%) | 2 (2.8%) | 0 (0%) |
| Number of subjects |  | 33 | 72<sup>a</sup> | 59 |
| Samples<br>by<br>timepoint | Uninfected Cross-sectional (N [%]) | - | - | 59 (100%) |
|  | Day 0 (N [%]) | 33 (14.6%) | 71 (74.0%) | - |
|  | Week 2 (N [%]) | 31 (13.7%) | 13 (13.5%) | - |
|  | Week4 (N [%]) | 32 (14.2%) | 12 (12.5%) | - |
|  | Month 3 (N [%]) | 30 (13.3%) | - | - |
|  | Month 6 (N [%]) | 12 (5.3%) | - | - |
|  | Month 12 (N [%]) | 32 (14.2%) | - | - |
|  | Month 18 (N [%]) | 29 (12.8%) | - | - |
|  | Month 24 (N [%]) | 23 (10.2%) | - | - |
|  | Reinfection (N<br>[%]) | 4 (1.8%) | - | - |
| <sup>a</sup>This group comprises 16 longitudinal subjects, each sampled at a minimum of two timepoints, and 56 cross-sectional subjects. |  |  |  |  |

### Antibodies to AgSAP and AgTRIO are higher in malaria endemic areas

Human antibody responses to mosquito salivary antigens AgSAP and AgTRIO were significantly higher among people with acute and recent infection from both Kédougou and Thiès compared to the HCW cohort (for AgSAP: compared to Kédougou acute and recent and Thiès recent, all p-values < 0.0001, compared to Thiès acute, p-value = 0.0136; for AgTRIO: all p-values < 0.001), and additionally AgTRIO antibodies were higher among Thiès non-transmission season samples compared to HCW (p=0.0007) (Figure 1A-B). Antibody levels against these two proteins were also higher for some uninfected people in these malaria-endemic areas compared to the negative HCW cohort. Specifically, antibodies to AgSAP were higher in uninfected people from Kédougou during the malaria transmission season (p-value = 0.0245), and antibodies to AgTRIO were higher in uninfected individuals for both Thiès and Kédougou during the malaria transmission season (p-values = 0.0064 and <0.0001, respectively).

**Figure 1.**
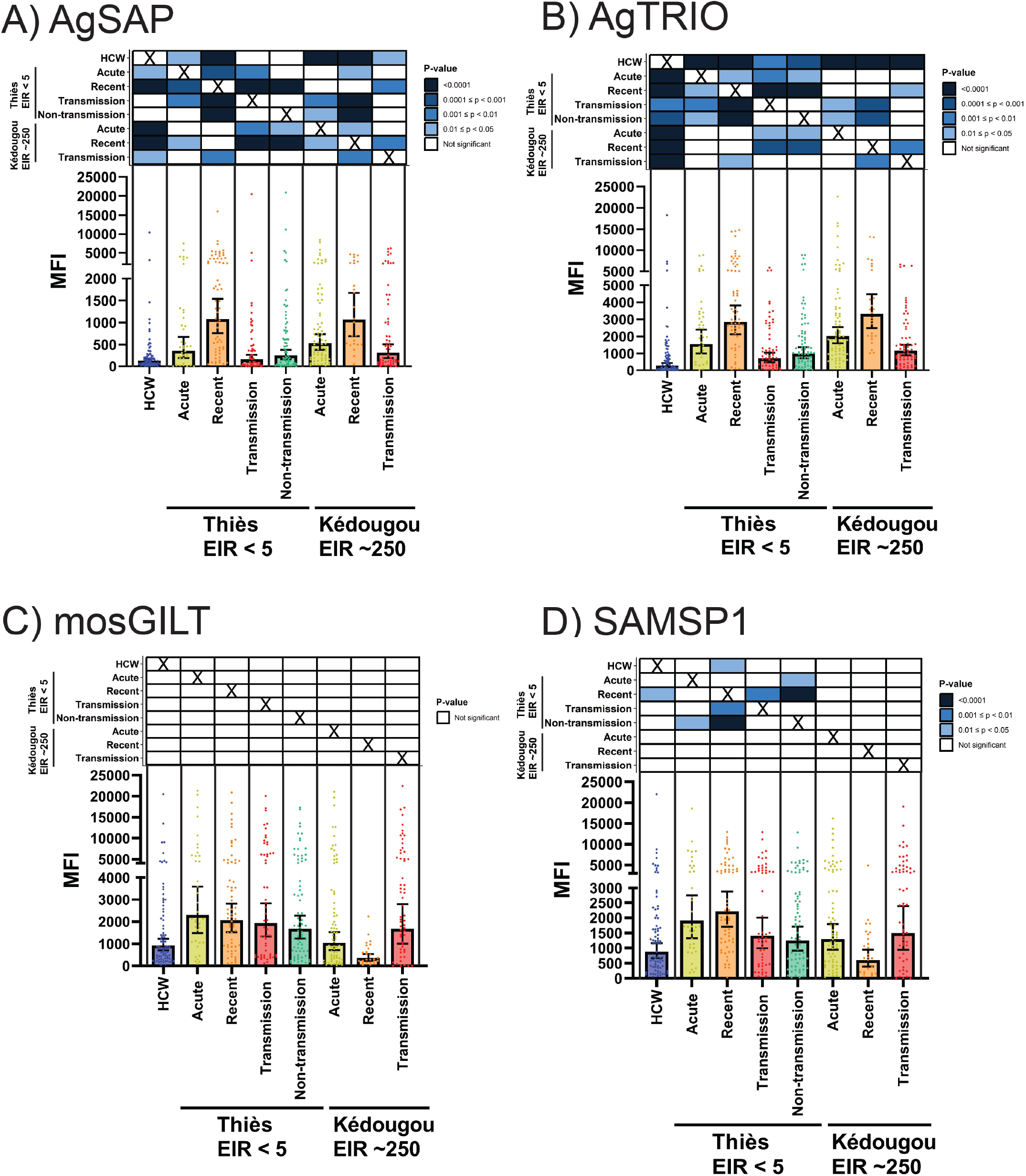
MFI differences between cohorts for IgG responses to the mosquito salivary antigens for A) AgSAP B) AgTRIO C) mosGILT and D) SAMSP1 with geometric means shown by bars, 95% confidence intervals shown by whiskers, and individual responses shown by dots. Cohorts are split by malaria transmission (Thiès is a low transmission area with an EIR <5 infectious bites per person per year and Kédougou is a moderate malaria transmission area with an EIR ∼250 infectious bites per person per year) and time relative to malaria infection (acute are samples from the time of malaria infection, recent are samples collected 2-4 weeks after malaria infection, transmission are samples collected from uninfected individuals during the malaria transmission season, and non-transmission are samples collected from uninfected individuals outside of the malaria transmission season). HCW is a cohort of health care workers from the US that have not been exposed to malaria infected mosquitoes. Heatmaps above each bar plot show significance differences between groups of liner mixed models of log transformed MFI of post-hoc pairwise comparisons adjusting p-values with Tukey’s method. X’s indicate comparisons were not performed.

The HCW cohort had a relatively high reactivity for the mosGILT and SAMSP1 proteins, resulting in no significant differences between HCW and uninfected individuals in the transmission season in either Thiès (mosGILT: p-value = 0.237; SAMSP1: p-value = 0.620, Figure 1C&D) or Kédougou (mosGILT: p-value = 0.328; SAMSP1: p-value = 0.342). The only significant difference was for SAMSP1 HCW cohort compared to Thiès recent infection cohort (p-value = 0.0182).

### Antibody responses to all proteins did not differ by transmission season nor intensity

We sought to determine whether responses could distinguish between seasons of high and low transmission and between regions of different malaria endemicity. None of the salivary antigens probed distinguished any significant differences between antibody responses from uninfected people in the transmission and non-transmission seasons for Thiès (Figure 1), the only cohort where it was possible to assess this comparison. There were also no differences in antibody responses to any of the antigens tested when comparing equivalent cohorts in the low transmission area of Thiès and the moderate transmission area of Kédougou (i.e. comparing categories of acute infection, recent infection, and uninfected transmission season samples between Thiès and Kédougou, Figure 1).

### Antibodies to AgSAP and AgTRIO were associated with recent infectious mosquito exposure

We next evaluated whether responses were associated with recent infectious mosquito bites. Immune reactivity to AgSAP and AgTRIO distinguished between cohorts with and without recent infectious mosquito exposure. For AgSAP, among people in Thiès, people with acute or recent (2-4 wks post diagnosis) malaria infection had significantly higher reactivity than uninfected people in the transmission season (p-value = 0.0078 < 0.0001, respectively; Figure 1A). The same was true for AgTRIO (p-values < 0.001; Figure 1B). However, among people in Kédougou, only those with recent infection had significantly higher AgSAP and AgTRIO reactivity than those uninfected from the transmission season for those (p-value = 0.0011 and 0.0049, respectively); there were no significant differences between those with acute infection compared to uninfected people in the transmission season.

For SAMSP1, recent infection in Thiès also increased IgG reactivity compared to uninfected individuals in the transmission season in Thiès (p-value = 0.0043), but other comparisons were not significant (Figure 1D). For mosGILT, reactivity among people with acute and recent infection were not significantly different than uninfected people in the transmission season (Figure 1C).

### Longitudinally, antibodies to AgSAP and AgTRIO peaked at 2-4 weeks post infection and decreased by 3 months after infection

Peak AgSAP reactivity in the Thiès cohort was among individuals who had malaria infection 2 weeks prior (significantly higher reactivity than all timepoints other than Week 4 and reinfection, p-values <0.001) and 4 weeks prior (significantly higher reactivity than at timepoints at 3, 12, 18, and 24 months after infection, p-values <0.001; Figure 2A & Table S2). Among the Kédougou cohort, reactivity 2 weeks and 4 weeks after infection were also significantly higher than at acute infection (p-values = 0.01 and 0.005, respectively).

**Figure 2.**
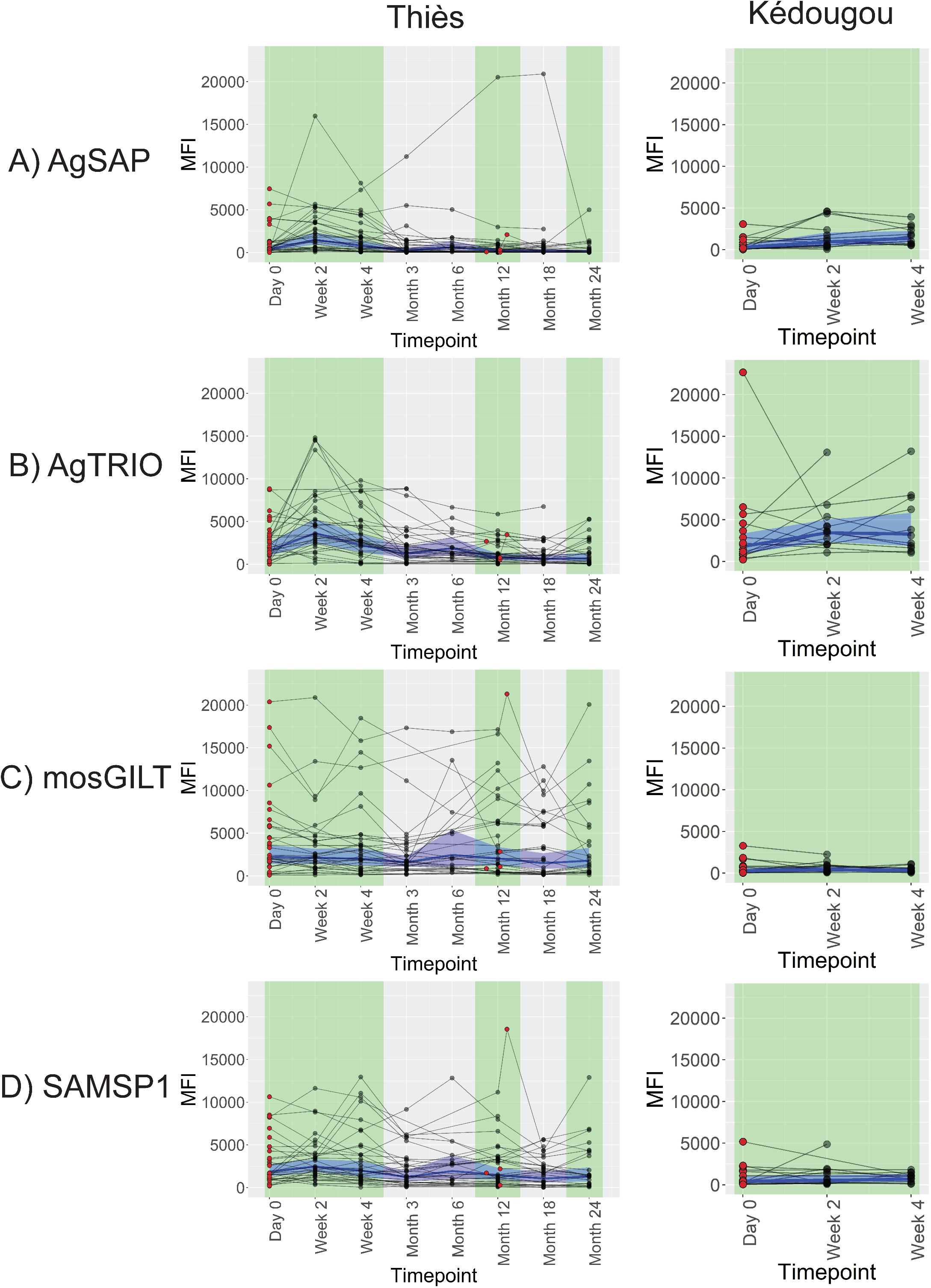
IgG MFI shown longitudinally by individual from Thiès and Kédougou for each mosquito salivary antigen A) AgSAP B) AgTRIO C) mosGILT and D) SAMSP1. Timepoints with infection shown in red (Day 0 and Reinfection for Thiès, and Day 0 for Kédougou), and timepoints without infection shown in black. Malaria transmission seasons shown in green. Blue line shows the geometric mean at each timepoint, and light blue shows the 95% confidence intervals for the geometric mean (calculated excluding reinfection timepoints for Thiès).

Similar to AgSAP, the highest AgTRIO antibody responses in the Thiés cohort were among individuals who had malaria infection 2 weeks prior (significantly higher reactivity than all timepoints other than Week 4 and reinfection, p-values <0.01) and 4 weeks prior (significantly higher reactivity than timepoints at 3 months [p-value = 0.03], 12, 18, and 24 months after infection, all p-values <0.0001; Figure 2B). However, while AgTRIO reactivity among the Kédougou cohort was also highest at 2 weeks and 4 weeks after infection, this was not significantly higher than at acute infection (p-value = 0.08 compared to Week 2, and p-value = 0.09 compared to Week 4).

For mosGILT, there were no significant antibody reactivity differences between timepoints in either the Thiès or Kédougou cohorts (Figure 2C). For SAMSP1 antibodies, in the Thiès cohort, reactivity among individuals who had malaria 2 weeks prior was significantly higher than reactivity among samples taken 3 months (p-value= 0.0008), 6 months (p-value = 0.049), 12 months (p-value = 0.02), 18 months (p-value <0.0001), and 24 months (p-value = 0.005) after infection (Figure 2D). There were no significant differences between timepoints in the Kédougou cohort for SAMSP1 reactivity.

### Using cutoffs for seropositivity, AgSAP had a high sensitivity and AgTRIO had a high specificity

AgSAP did not demonstrate a bimodal distribution and as such, using the negative HCW cohort, the positive cutoff value for AgSAP was an MFI of 1645 (calculated from mean + 2 standard deviations of log transformed results: Figure S2A). The specificity was 97.4% among the HCW cohort (75 of 77, Figure 3). The sensitivity of AgSAP was 18.9% (7 of 37) and 22.5% (16 of 71)among individuals in Thiès and Kédougou, respectively, with acute infection. The sensitivity of AgSAP was 47.6% (30 of 63) and 44.0% (11 of 25) among individuals in Thiès and Kédougou, respectively, with recent infection (2-4 weeks prior).

**Figure 3.**
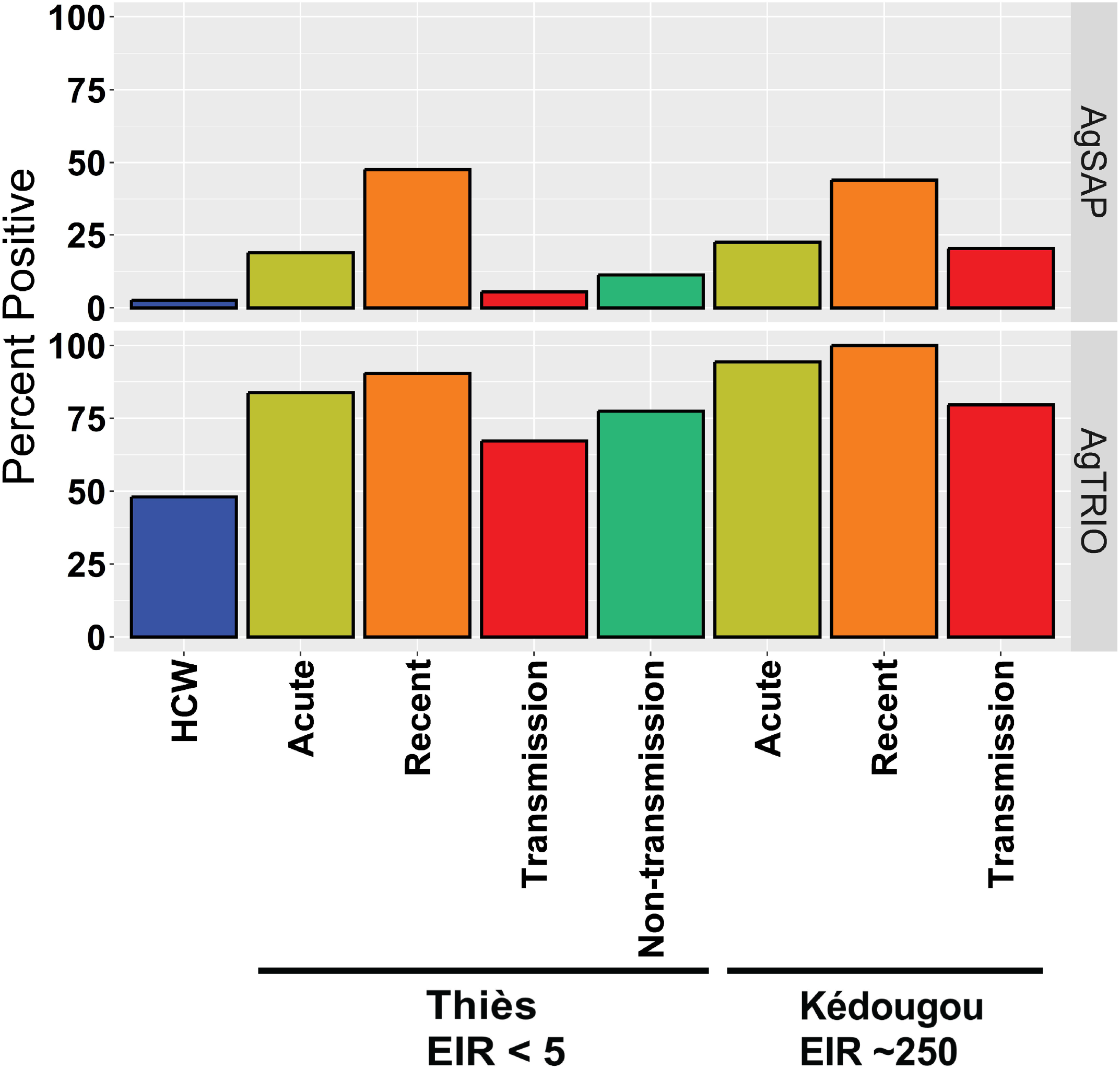
Seropositivity for each cohort for mosquito salivary antigens AgSAP and AgTRIO. Cohorts are split by malaria transmission (Thiès is a low transmission area with an EIR <5 infectious bites per person per year and Kédougou has a moderate malaria transmission with an EIR ∼250 infectious bites per person per year) and time relative to malaria infection (acute are samples from the time of malaria infection, recent are samples collected 2-4 weeks after infection, transmission are samples collected from uninfected individuals during the transmission season, and non-transmission are samples collected from uninfected individuals outside of the transmission season). HCW is a cohort of health care workers from the US that are not expected to have exposure to malaria infected mosquitoes.

In contrast, AgTRIO MFI values exhibited a mildly bimodal distribution so using the FMM from a bimodal distribution, the positive cutoff value for AgTRIO was MFI of 500 (Figure S2B). Results from the HCW cohort for AgTRIO were too high to obtain a usable cutoff. Specificity for the HCW cohort was 51.9% (40 of 77). The sensitivity of AgTRIO was 83.8% (31 of 37) and 94.4% (67 of 71) among individuals in Thiès and Kédougou, respectively, with acute infection. The sensitivity of AgTRIO was 90.5% (57 of 63) and 100% (25 of 25) among individuals in Thiès and Kédougou, respectively, with recent infection.

### Groups with recent infection had highest rates of seropositivity

AgSAP seropositivity was higher in samples 2 weeks after malaria infection compared to HCW for individuals from both Thiès (p-value < 0.0001) and Kédougou (p-value = 0.0033), as well as when comparing uninfected individuals from Kédougou during the transmission season to HCW (p-value = 0.0043). Similarly, compared to HCW, seropositivity for AgTRIO was higher in samples 2 weeks after malaria infection in people from both Thiès and Kédougou (p-values < 0.001 and = 0.0017, respectively), as well as in uninfected individuals from Kédougou during the transmission season (p-value = 0.0013).

### Responses to AgSAP and AgTRIO were correlated

Of 458 total samples tested by multiplex,, 87 samples were positive for both AgSAP and AgTRIO, and 100 were negative by both AgSAP and AgTRIO using the cutoffs calculated for positivity. 269 samples were positive by AgTRIO and negative by AgSAP cutoffs, and 2 samples were positive by AgSAP and negative by AgTRIO cutoffs. MFI results of AgSAP and AgTRIO had a Spearman correlation of rho = 0.636 (p-value < 0.0001, Figure S3).

### IgG responses to mosquito salivary antigens as measured by sample type and testing platform were moderately correlated

For a subset of Kédougou samples (n = 58), both plasma and DBS samples were tested on the multiplex platform to compare results of different sample types. Among these matched samples, AgSAP had the highest correlation between results for plasma and DBS (Pearson correlation r = 0.492, p-value < 0.0001, Figure S4). The next highest correlation involved SAMSP1 (Pearson correlation r = 0.388, p-value = 0.003) and mosGILT (Pearson correlation r = 0.383, p-value = 0.003), and then AgTRIO (Pearson correlation r = 0.229, p-value = 0.08).

A subset of 120 samples from Thiès and 30 HCW samples were also tested by both ELISA and multiplex to compare results between testing platforms (Figure 4). Samples tested by ELISA and multiplex assays were moderately correlated for both the AgSAP protein (rho = 0.394, p-value <0.0001) and the AgTRIO protein (rho = 0.330, p-value <0.0001). However, HCW samples tested by ELISA showed high background for both AgSAP (median OD = 0.507) and AgTRIO (median OD = 0.424) relative to Thiès samples tested (AgSAP median OD = 0.530, AgTRIO median OD = 0.306).

**Figure 4.**
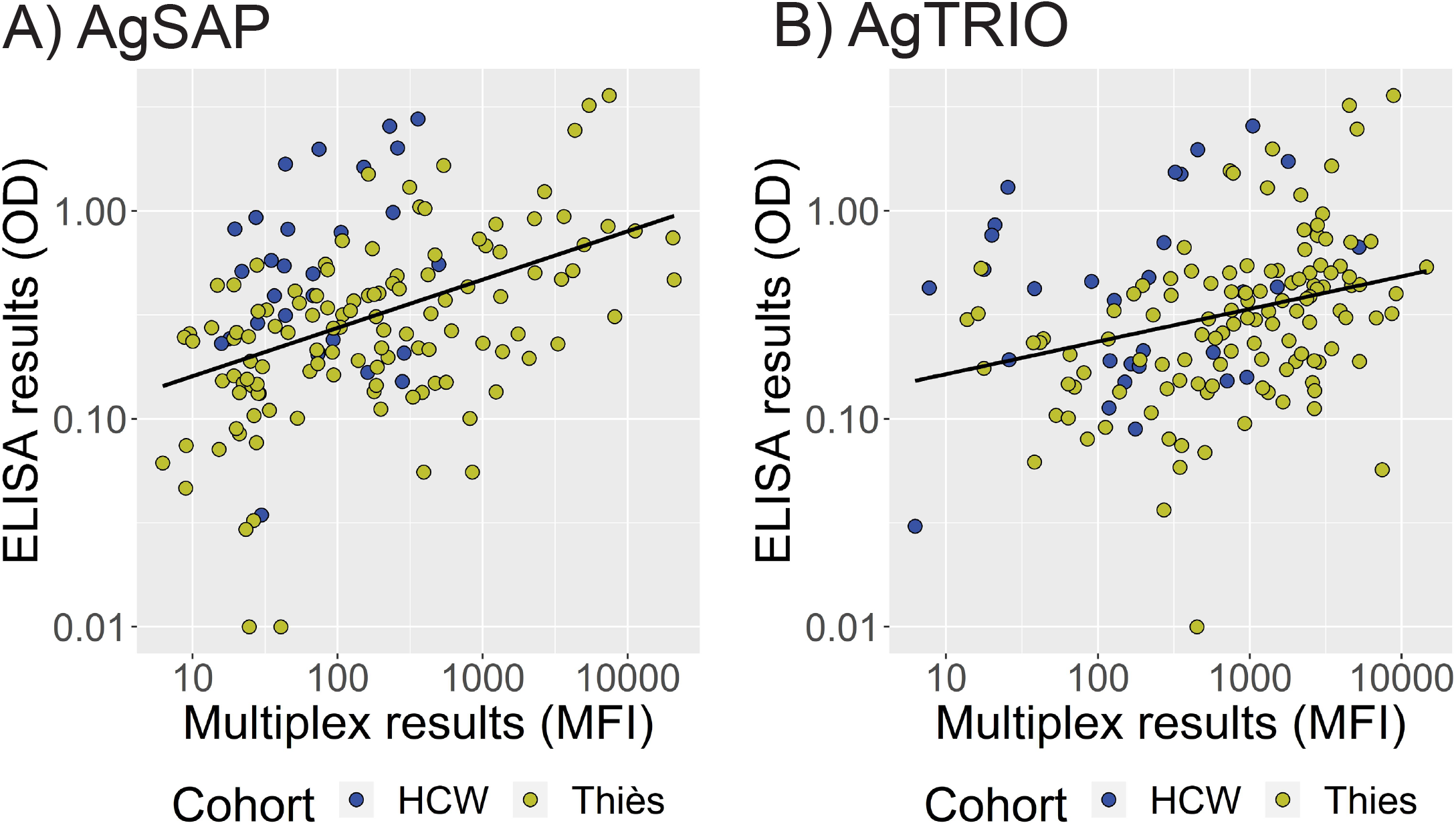
Multiplex and ELISA comparisons for A) AgSAP and B) AgTRIO with HCW samples shown in blue and Thiès samples shown in yellow. Two samples for AgSAP and one for AgTRIO with values between 0.02 and 0 were set to 0.01 to allow them to be shown on a log scale.

## Discussion

### Advantages of serological multiplex approach

While serological multiplex approaches have previously been used with *P. falciparum* antigens to measure malaria prevalence and time from last infection[3-5], here we determine that serological responses to MSAs can also effectively indicate exposure to infectious mosquitoes. This could be an efficient alternative to EIR as a measure of *P. falciparum* transmission intensity from mosquitoes to humans. Serological responses to MSAs could be preferable to EIR as an estimate of *P. falciparum* transmission from mosquitoes to humans, since EIR estimates are limited by sampling issues (such as seasonal variation), precision (such as variation in measuring human biting rate and contribution of indoor/outdoor biting), and accuracy (such as spatial and temporal heterogeneity in vector density)[2]. Serological measurements are a more direct measure of the effect of mosquito bites on humans, and therefore do not have the same issues. Among the four MSAs tested, human IgG responses to AgSAP and AgTRIO were the best indicators of recent exposure to infectious mosquitos.

### Higher IgG reactivity to AgSAP and AgTRIO is associated with malaria endemicity

We show here that reactivity to AgSAP is often higher in populations in malaria endemic areas than a non-endemic area, which is consistent with a previous study[21]. Here, this was true for the moderate transmission area of Kédougou compared to healthy controls in a non-endemic area (p-value = 0.02). However, AgSAP reactivity did not discriminate between populations in areas of different malaria endemicity (all p-values > 0.05).

Similarly, AgTRIO responses were higher in populations in malaria endemic areas than a non-endemic area. We found that uninfected individuals in both low and moderate transmission areas had higher AgTRIO reactivity than healthy controls in a non-endemic area (p-value = 0.0064 for Thiès and p-value <0.0001 for Kédougou). Although a previous study found that AgTRIO responses in individuals from a malaria endemic region were non-significantly higher than in individuals from a non-endemic area[23], our study could have been appropriately powered to observe the differences. Like AgSAP, reactivity to AgTRIO could not discriminate between populations in areas of different malaria endemicity (all p-values > 0.05)

Although mosGILT was found to be directly associated with *Plasmodium* sporozoites during transmission[20], serological responses to mosGILT did not show clear patterns in relation to infectious mosquitoes exposure. There were no significant differences between groups or timepoints, so this study does not provide evidence for utilizing mosGILT in malaria transmission surveillance. The high reactivity of mosGILT for the negative HCW cohort limited our ability to distinguish between populations. Similarly, SAMSP1 reactivity did not differ between populations in malaria endemic areas and a non-endemic area. We found healthy controls in a non-malaria endemic area did not have significantly different SAMSP1 reactivity than uninfected individuals in Thiès (all p-values > 0.05). This contrasts with a previous study, which found that individuals in a malaria-endemic area in Senegal had higher IgG reactivity to SAMSP1 compared to a group of individuals living in a non-endemic area in France[22]. However, as with mosGILT, the high reactivity of SAMSP1 in our HCW cohort limits our ability to observe differences between populations, which could account for these different findings.

### Mosquito salivary proteins are not biomarkers of general mosquito exposure or malaria transmission season alone, in the absence of infection

Since there were no significant differences between uninfected people in the transmission and non-transmission seasons for Thiès (comparison not available for Kédougou), there is no evidence that these mosquito salivary proteins can determine seasonal differences in malaria exposure intensity. Furthermore, there were no significant differences between equivalent cohorts in our low and moderate transmission areas, showing no evidence of differences between populations exposed to variable malaria transmission intensity.

### AgSAP and AgTRIO are promising indicators of recent or infectious mosquito exposure

In a low transmission area, where infectious mosquito bites are less common, IgG responses to AgSAP peak after infectious mosquito exposure and decline by 3 months after infection. Since AgSAP is upregulated in infectious mosquitoes[21], a bite from an infectious mosquito could induce a greater serological response than a bite from an uninfected mosquito. AgSAP reactivity was highest among individuals with recent infection, suggesting that AgSAP serological responses peak 2-4 weeks after clinical malaria, which is itself at least 7-10 days after an infectious bite[36]. In both the low transmission area of Thiès and the moderate transmission area of Kédougou, AgSAP responses during recent infection were significantly higher than during acute infection (p-value = 0.0002 for Thiès and p-value = 0.02 for Kédougou). AgSAP responses decline after recent infection and are significantly lower 3 months after initial infection (p-value = 0.0001 in Thiès comparing timepoints 4 weeks and 3 months after infection; not tested in Kédougou), suggesting the kinetics of AgSAP build a couple weeks after infection, and then decline relatively quickly.

The AgTRIO trends were similar to those for AgSAP. IgG responses to AgTRIO also peak in the weeks after exposure to an infectious mosquito and decline by 3 months after infection. Like AgSAP, AgTRIO is upregulated in *Plasmodium* infected mosquitoes[23], so a bite from an infectious mosquito could induce a greater serological response than a bite from an uninfected mosquito. Reactivity to AgTRIO was also highest among individuals with a recent infection, suggesting that AgTRIO serological responses peak 2-4 weeks after clinical malaria. AgTRIO reactivity was significantly higher 2 weeks after infection than at the time of acute infection in the Thiès cohort (p-value = 0.0018). Like AgSAP, AgTRIO responses significantly declined 3 months after initial infection (p-value = 0.024 in Thiès comparing timepoints 4 weeks and 3 months after infection; not tested in Kédougou), suggesting that like AgSAP, the kinetics of AgTRIO build a couple weeks after infection, and then decline relatively quickly.

SAMSP1 IgG responses do seem to increase in individuals with acute and recent malaria infection relative to reactivity outside of the transmission season (in the Thiès cohort, acute infection p-value = 0.029; and recent infection p-value <0.0001). In Thiès, recent infection also increased reactivity compared to uninfected individuals in the transmission season (p-value = 0.0043) and compared to healthy individuals from a non-malaria endemic area (p-value = 0.018). This also indicates that SAMSP1 reactivity may peak 2-4 weeks after infection and decrease by 3 months after infection, although differences were not observed in Kédougou.

### Seropositivity could show differences between populations

For AgSAP and AgTRIO, cutoffs for positivity allow for moderate discrimination among cohorts. However, the greater benefit of these proteins may be to discriminate among changes in populations rather than at the individual level, for example, before and after an intervention is rolled out in a community. Bimodal distributions can be an effective way to determine cutoffs, and determining a cutoff for AgTRIO using this method may benefit from the inclusion of more samples[33]. For the cutoffs used here, AgSAP had a relatively low sensitivity and a high specificity. Conversely, AgTRIO had high sensitivity and a lower specificity. Both AgSAP and AgTRIO had higher sensitivity among individuals with recent infection than acute infection.

As multiple *P. falciparum* antigens have been used to estimate malaria prevalence[4, 5, 37], we investigated whether AgSAP and AgTRIO results together could increase sensitivity and specificity. However, using our established cutoffs, very few samples were positive by AgSAP and negative by AgTRIO (2 of 458). Additionally, results between AgSAP and AgTRIO were highly correlated (Spearman rho = 0.636), and kinetics of responses appear similar, with responses to both antigens peaking between 2-4 weeks after infection. Therefore, using responses to these antigens together does not improve our ability to discriminate between samples with and without recent malaria infection.

### Reactivity for different sample types should be tested further

Pearson correlations for a subset of samples tested from plasma and DBS were moderately correlated for AgSAP, mosGILT, and SAMSP1 proteins (Pearson correlation r between 0.383 and 0.492) and weakly correlated for AgTRIO (Pearson correlation r = 0.229). Since DBS are often used for population-level surveillance because of reduced cost and collection burden versus plasma, DBS should be further tested to determine whether the patterns observed here for plasma also hold for DBS.

### Reactivity for AgSAP and AgTRIO tested by ELISA and multiplex assays are moderately correlated

For both AgSAP and AgTRIO, samples tested by ELISA and multiplexed assay were moderately correlated, suggesting it may be possible to use ELISA for surveillance using these proteins where multiplex approaches are not available. However, the high background for AgSAP and AgTRIO ELISA results among negative samples from the HCW cohort should be investigated further to ensure that ELISAs can confidently discriminate between samples from malaria endemic and non-endemic areas before these proteins are used for population-level surveillance.

### Cohorts in context

This study tested samples from two different regions of Senegal: Thiès, a low transmission area with an EIR of <5 infectious bites per person per year[34, 38, 39], and Kédougou, a moderate transmission area with an EIR ∼250[35]. The low malaria transmission in Thiès has been tracked over time[40, 41], and in the longitudinal cohort tested here, only four of seventy individuals had malaria reinfections over two years of follow-up. Conversely, since malaria transmission is higher in Kédougou, we expect this cohort to have been exposed to infectious mosquitoes more frequently than the Thiès cohort. This could affect immune responses to MSAs between these areas, particularly antigens that are upregulated in *Plasmodium*-infected mosquitoes. Therefore, individuals from Kédougou may be more likely to get a ‘boosting’ effect from exposure to infected mosquitoes.

As such, the longitudinal changes in responses from Thiès may demonstrate a more direct indicator of responses after exposure to antigens from a *Plasmodium*-infected mosquito. The longitudinal data from Thiès clearly shows peaks in AgSAP and AgTRIO responses 2-4 weeks after subjects had symptomatic malaria, and then tend to reduce for the subsequent two years of follow-up. The longitudinal data from the moderate-transmission area of Kédougou likewise shows high levels of reactivity in AgSAP and AgTRIO up to four weeks after symptomatic malaria, though without longer follow-up we cannot compare longer term kinetics of antibody responses in Kédougou to those in Thiès.

### Limitations

We tested samples from a low and moderate transmission area of Senegal, but we cannot exclude the possibility that serological results might be different in an extremely high transmission area. Although the kinetics of AgSAP and AgTRIO observed in this study suggested that serological responses diminish within a couple of months, a person in a high transmission area could be exposed to infectious mosquitoes more frequently. Our Kédougou cohorts were meant to model high transmission, but they are still relatively lower compared to other regions with intense, hyperendemic transmission. In a high transmission area with more infectious mosquito exposure, serological responses might remain high among the population, which could obscure changes in malaria transmission. Additionally, we cannot distinguish between infected mosquito bites and extremely recent exposure to mosquito bites without evaluating humans known to be bitten by uninfected mosquitoes recently but who did not develop infection.

Different geographical regions with a different distribution of mosquito species could also affect serological results. Cross-reactivity across *Anopheles* species has yet to be empirically tested.

## Conclusions

This is a promising first study investigating human reactivity to four novel MSAs in low and moderate malaria transmission areas. These salivary antigens represent potential complimentary antigens to SG6, an MSA that is a marker of exposure to mosquito bites, and *P. falciparum* antigens that are markers of malaria parasite exposure to different stages of the parasite life cycle, as antigens that uniquely represent potential markers of recent exposure to infectious mosquito bites. This study showed AgSAP and AgTRIO to be promising markers for exposure to infectious mosquito bites in low and moderate transmission areas. Further studies could investigate their utility in geographical areas with higher malaria transmission intensities. The multiplex approach used here is a flexible method that allows for the possibility of adding additional markers, which is encouraging for testing these antigens further in high transmission areas and more geographical regions. This could be particularly useful in regions already using a multiplex approach for disease surveillance in order to efficiently measure population-level changes in *P. falciparum* exposure, especially in populations before and after implementation of vector control interventions.

## Data Availability

All data produced in the present study are available upon reasonable request to the authors and additionally are available online at https://doi.org/10.5061/dryad.cz8w9gjbj.

## Acknowledgements

We acknowledge all study participants and communities in Thiès, Senegal and Kédougou, Senegal for their participation in this project. We further acknowledge the Health Care Workers from Yale New Haven Hospital who participated in this project. We wish to thank Alioune Wade for assistance with sample processing in Kédougou. We acknowledge Dyann F. Wirth for her mentorship on the Fogarty International Center K01, and continued support.

## Author Contributions

AKB, YC, and EF conceived and directed the project, SL and MG performed the experiments, HR assisted with bead coupling, SL analyzed the data, MS, ABD, YD, AMM, KH, SDS, AW, MNP, LGT, AB, NG, AM, DFW, DN, SP, AK, and AKB performed data acquisition, cohort initiation, follow-up and maintenance. AKB, MG and SL wrote the manuscript. All authors have read, provided feedback, and approved the submitted version of the manuscript.

## Data accessibility

Data associated with this manuscript can be found at: DOI: 10.5061/dryad.cz8w9gjbj

## Figure Legends

**Table S1:** MFI Geometric mean with lower and upper 95% confidence intervals for all cohort groups and proteins

**Table S2:** MFI Geometric mean with lower and upper 95% confidence intervals for all cohort-timepoints and proteins

**Figure S1**. Sizes of the proteins, as verified by SDS-Page gel, were found to be: 70kDa for AgSAP, 42 kDa for AgTRIO, 30 kDa for mosGILT, and 45 kDa for SAMPS1.

**Figure S2**. Distribution of A) AgSAP and B) AgTRIO MFI multiplex results for all plasma samples tested. Cutoffs for positivity are 1645 for AgSAP and 500 for AgTRIO, as indicated by dashed lines

**Figure S3**. Scatterplot of results of AgSAP and AgTRIO. Vertical dashed line shows AgSAP cutoff for positivity (MFI of 1645), and horizontal dashed line shows AgTRIO cutoff for positivity (MFI of 500).

**Figure S4**. Scatterplots of log MFI results of matched samples of plasma and dried blood spots for the four mosquito salivary antigens

