## Supplemental Methods for "Two mosquito salivary antigens demonstrate promise as biomarkers of recent exposure to *P. falciparum* infected mosquito bites"

**Ethical approvals:** *HCW:* This study was approved by the Yale Human Investigation Committee, protocol #2000027690. Samples were collected with informed consent and in accordance with Yale IRB approval.

*Thiès:* This study was approved by the National Ethics Committee of the Ministry of Health in Senegal (Protocol SEN 14/49), the Institutional Review Board of the Harvard T.H. Chan School of Public Health (IRB 14-2830), and the Human Investigation Committee of Yale University (Protocol 2000023287). Samples were collected with informed consent and in accordance with all ethical requirements of the aforementioned entities.

*Kédougou (2019 and 2023):* These studies were approved by the National Ethics Committee of Senegal (CNERS) (Protocols SEN19/37 and SEN23/09) and the Institutional Review Board of the Yale School of Public Health (Protocols 2000025417 and 2000035379*)*. Research was performed in accordance with relevant guidelines and regulations, and informed consent was obtained from all participants and/or their legal guardians. Samples from the Kédougou cross-sectional cohort were collected as part of Institut Pasteur de Dakar surveillance of febrile illness.

**Cohorts:** *HCW:* The cohort of health care workers (HCWs) from Yale New Haven Hospital in New Haven, CT were prospectively followed as part of the Yale Implementing Medical and Public Health Action Against Coronavirus CT (IMPACT) study after testing negative for SARS-CoV-2. This cohort includes 77 HCWs who contributed serum between April 13, 2020 and April 23, 2020, having tested negative for SARS-CoV-2 infection by RT-qPCR since the study start. They are not expected to have immune reactivity to these mosquito salivary proteins.

*Thiès:* Samples from the low-transmission area of Thiès, Senegal (70 km West of Dakar) come from individuals who presented through passive case detection at the Service de Lutte Anti Parasitaire clinic in 2015-2017 with malaria-like symptoms, tested positive via *P. falciparum*-specific HRP2/3 rapid diagnostic test (RDT), and had positive microscopy for *P. falciparum* monogenomic infection. Since individuals in the longitudinal cohort are enrolled through passive case detection and are *P. falciparum* positive on Day 0 ^30^, these individuals were likely bitten by *P. falciparum*-infected mosquitoes up to 10 days prior ^31^ . Subjects were followed for two years, with samples collected at Day 0, Week 2 and 4, and Months 3, 6, 12, 18, and 24, and if reinfected. This group is split into categories of: Acute infection (Day 0 and Reinfection), Recent infection (Weeks 2 and 4), Transmission season (Months 12 and 24); and Non-transmission season (Months 3, 6, and 18,given initial case detection occurred during the seasonal transmission season, which, in Senegal, is from July to December with an October peak).

*Kédougou (2019) Cross-sectional:* Samples from the moderate-transmission area of Kédougou, Senegal, located in Southeastern Senegal 710 km from Dakar, were collected in July 2019. Patients were recruited from five clinics in Kédougou. Eligibility criteria was fever (temperature ≥38°C) including in the past 24 hours. *P. falciparum* positivity was determined based on a *P. falciparum*-specific HRP2/3 RDT. Thin and thick blood smears confirmed monogenomic infection with *P. falciparum* by microscopy. All samples were collected during the transmission season at the time subjects presented to the clinic, and samples were split into those positive and negative by RDT. Those positive are in the Acute infection group; those negative are in the Transmission group (uninfected people from the transmission season).

*Kédougou (2023) Longitudinal:* Subjects were identified through passive case detection at two clinics in Kédougou. Blood and dried blood spots (DBS) were collected longitudinally, at time of presentation, and 2 and 4 weeks after. This group is split into Acute infection (Day 0) and Recent infection (Weeks 2 and 4).
