## Supplementary figures and images for "Two mosquito salivary antigens demonstrate promise as biomarkers of recent exposure to *P. falciparum* infected mosquito bites"

### Supplemental Figure 1

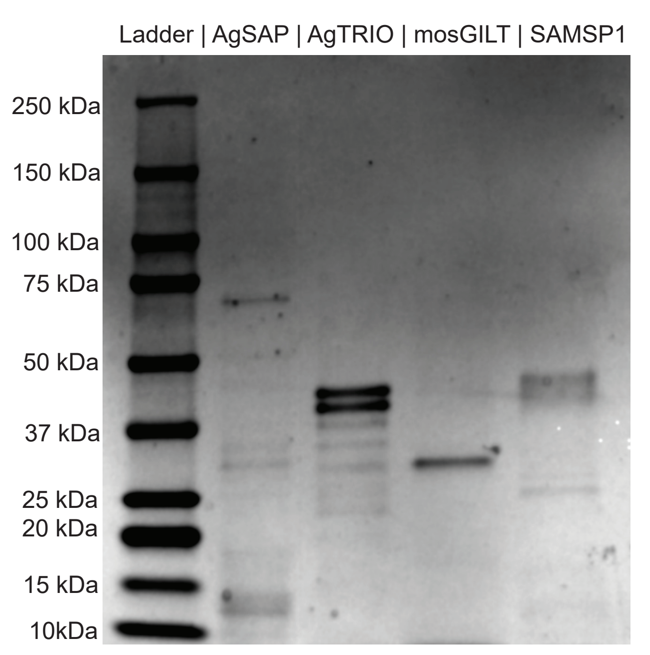

### Supplemental Figure 2

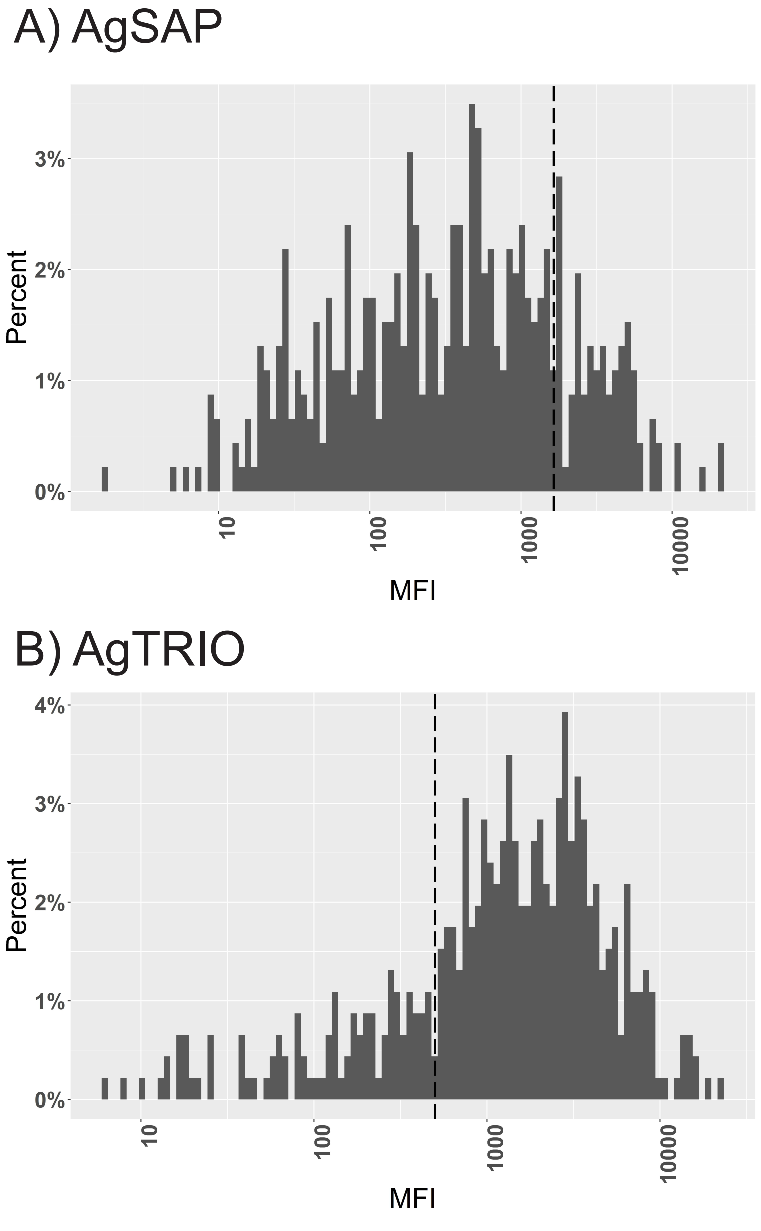

### Supplemental Figure 3

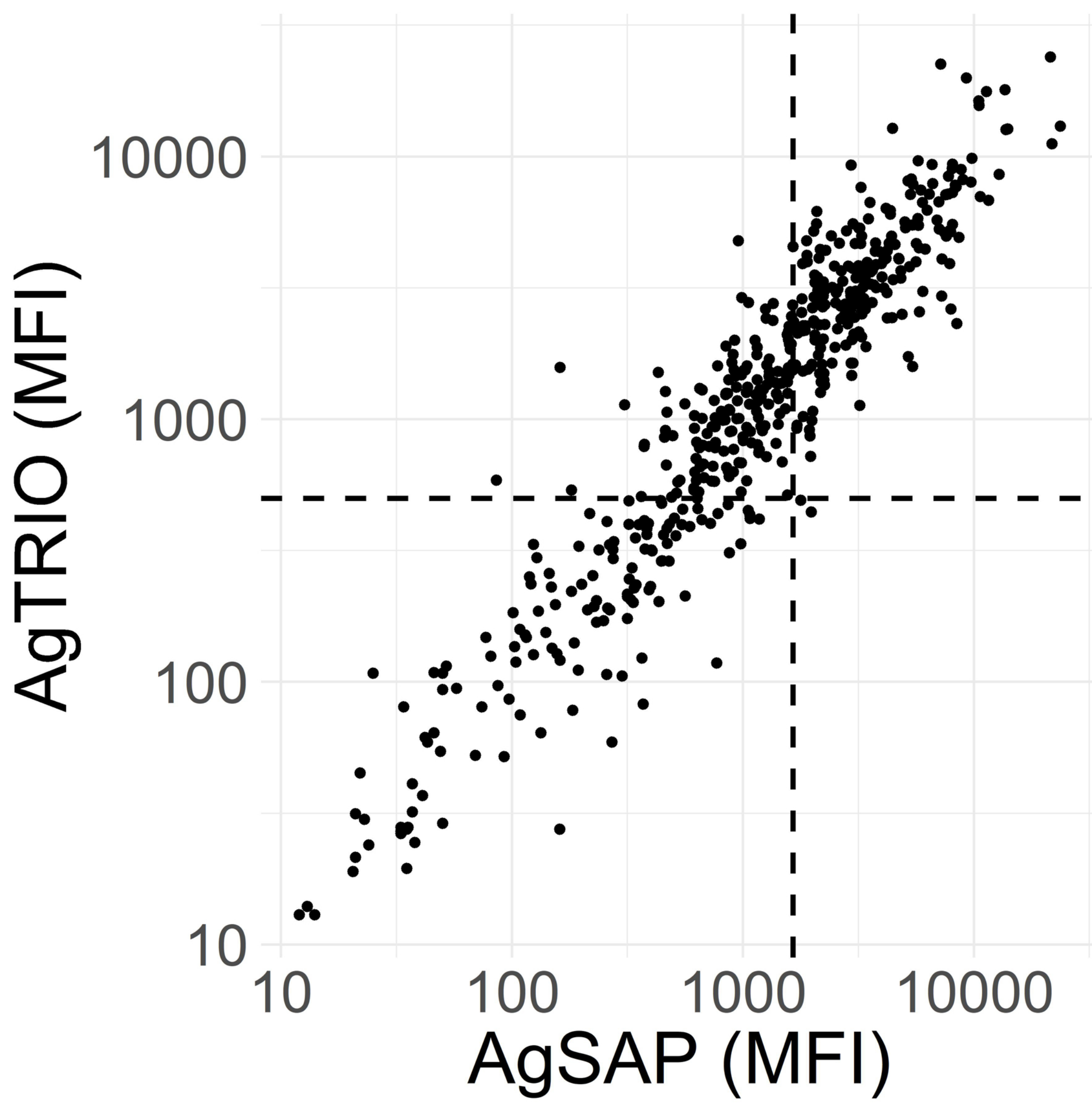

### Supplemental Figure 4

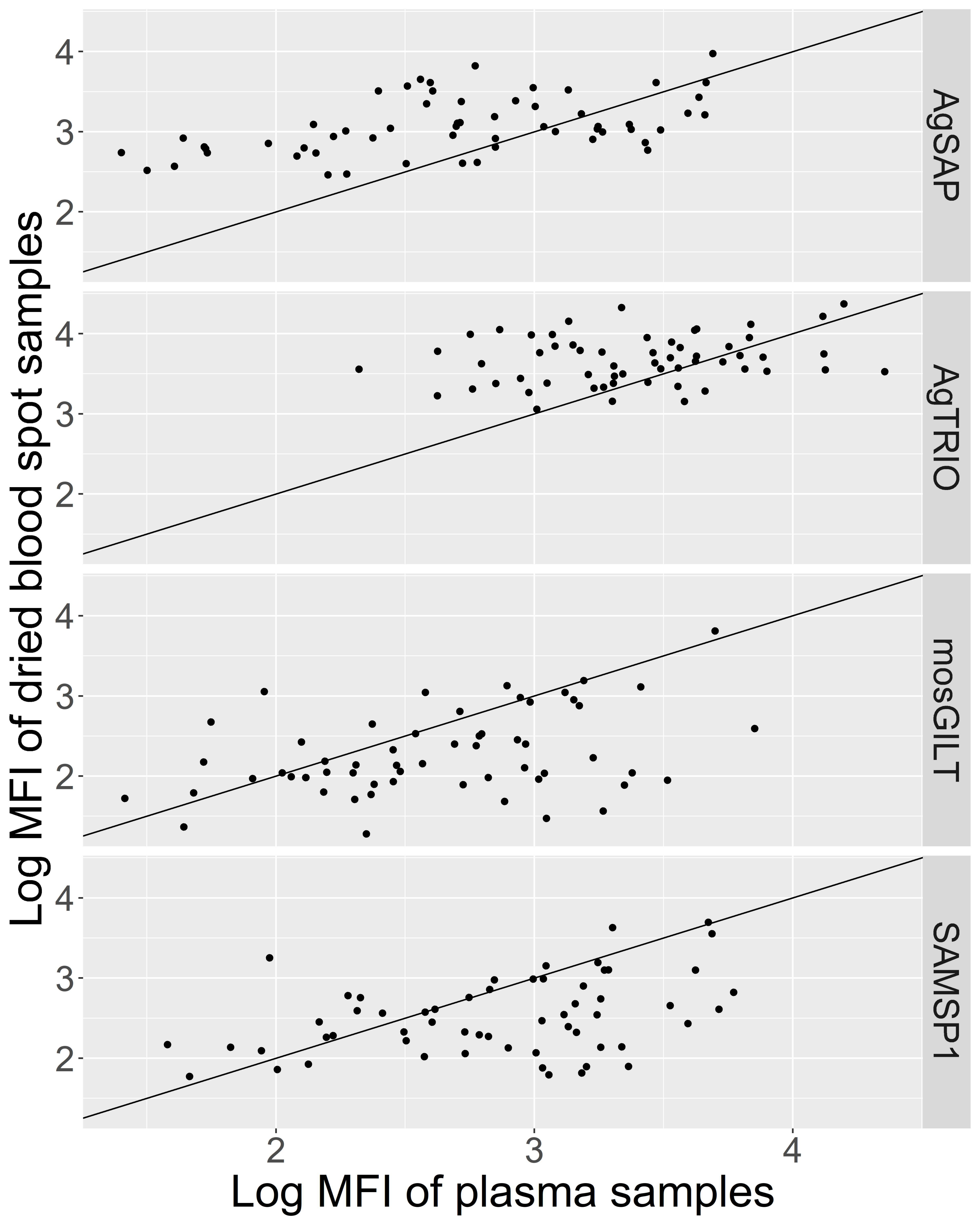
