## Supplemental Table 1 for "Two mosquito salivary antigens demonstrate promise as biomarkers of recent exposure to *P. falciparum* infected mosquito bites"

| Protein | Cohort group | Geometric mean (MFI) | Lower 95% CI for geometric mean (MFI) | Upper 95% CI for geometric mean (MFI) |
|---|---|---|---|---|
| AgSAP | Thiès recent infection | 1081 | 758 | 1542 |
| AgSAP | Kédougou recent infection | 1074 | 689 | 1674 |
| AgSAP | Kédougou acute infection | 531 | 384 | 735 |
| AgSAP | Thiès acute infection | 360 | 194 | 671 |
| AgSAP | Kédougou transmission season | 314 | 194 | 507 |
| AgSAP | Thiès Non-transmission Season | 248 | 163 | 378 |
| AgSAP | Thiès transmission season | 163 | 101 | 264 |
| AgSAP | HCW | 131 | 98 | 175 |
| AgTRIO | Kédougou recent infection | 3339 | 2490 | 4476 |
| AgTRIO | Thiès recent infection | 2847 | 2120 | 3823 |
| AgTRIO | Kédougou acute infection | 2023 | 1605 | 2551 |
| AgTRIO | Thiès acute infection | 1551 | 1001 | 2402 |
| AgTRIO | Kédougou transmission season | 1172 | 903 | 1521 |
| AgTRIO | Thiès Non-transmission Season | 998 | 725 | 1374 |
| AgTRIO | Thiès transmission season | 714 | 480 | 1063 |
| AgTRIO | HCW | 296 | 197 | 447 |
| SAMSP1 | Thiès recent infection | 2219 | 1707 | 2884 |
| SAMSP1 | Thiès acute infection | 1913 | 1327 | 2758 |
| SAMSP1 | Kédougou transmission season | 1501 | 944 | 2389 |
| SAMSP1 | Thiès transmission season | 1411 | 995 | 2002 |
| SAMSP1 | Kédougou acute infection | 1304 | 948 | 1794 |
| SAMSP1 | Thiès Non-transmission Season | 1252 | 918 | 1708 |
| SAMSP1 | HCW | 882 | 668 | 1166 |
| SAMSP1 | Kédougou recent infection | 605 | 385 | 950 |
| mosGILT | Thiès acute infection | 2302 | 1475 | 3593 |
| mosGILT | Thiès recent infection | 2068 | 1522 | 2810 |
| mosGILT | Thiès transmission season | 1935 | 1329 | 2817 |
| mosGILT | Kédougou transmission season | 1673 | 1004 | 2788 |
| mosGILT | Thiès Non-transmission Season | 1671 | 1235 | 2262 |
| mosGILT | Kédougou acute infection | 1044 | 714 | 1527 |
| mosGILT | HCW | 925 | 700 | 1222 |
| mosGILT | Kédougou recent infection | 352 | 233 | 531 |
