## Supplemental Table 2 for "Two mosquito salivary antigens demonstrate promise as biomarkers of recent exposure to *P. falciparum* infected mosquito bites"

| Protein | Cohort | Timepoint | Geometric mean (MFI) | Lower 95% CI for geometric mean (MFI) | Upper 95% CI for geometric mean (MFI) |
|---|---|---|---|---|---|
| AgSAP | Thiès | DAY 0 | 389 | 200 | 757 |
| AgSAP | Thiès | WEEK 2 | 1483 | 937 | 2348 |
| AgSAP | Thiès | WEEK 4 | 796 | 463 | 1368 |
| AgSAP | Thiès | MONTH 3 | 233 | 116 | 469 |
| AgSAP | Thiès | MONTH 6 | 626 | 291 | 1346 |
| AgSAP | Thiès | MONTH 12 | 192 | 101 | 365 |
| AgSAP | Thiès | MONTH 18 | 180 | 90 | 359 |
| AgSAP | Thiès | MONTH 24 | 131 | 61 | 280 |
| AgSAP | Thiès | REINFECTION | 191 | 10 | 3803 |
| AgSAP | Kédougou | DAY 0 | 285 | 135 | 600 |
| AgSAP | Kédougou | WEEK 2 | 928 | 419 | 2057 |
| AgSAP | Kédougou | WEEK 4 | 1367 | 832 | 2244 |
| AgTRIO | Thiès | DAY 0 | 1571 | 969 | 2547 |
| AgTRIO | Thiès | WEEK 2 | 3635 | 2502 | 5283 |
| AgTRIO | Thiès | WEEK 4 | 2247 | 1420 | 3554 |
| AgTRIO | Thiès | MONTH 3 | 1201 | 718 | 2009 |
| AgTRIO | Thiès | MONTH 6 | 1855 | 1108 | 3106 |
| AgTRIO | Thiès | MONTH 12 | 761 | 466 | 1242 |
| AgTRIO | Thiès | MONTH 18 | 638 | 376 | 1080 |
| AgTRIO | Thiès | MONTH 24 | 654 | 322 | 1330 |
| AgTRIO | Thiès | REINFECTION | 1393 | 322 | 6026 |
| AgTRIO | Kédougou | DAY 0 | 1793 | 960 | 3347 |
| AgTRIO | Kédougou | WEEK 2 | 3424 | 2334 | 5023 |
| AgTRIO | Kédougou | WEEK 4 | 3279 | 1851 | 5810 |
| mosGILT | Thiès | DAY 0 | 2254 | 1400 | 3627 |
| mosGILT | Thiès | WEEK 2 | 2119 | 1414 | 3177 |
| mosGILT | Thiès | WEEK 4 | 2020 | 1247 | 3269 |
| mosGILT | Thiès | MONTH 3 | 1599 | 1087 | 2353 |
| mosGILT | Thiès | MONTH 6 | 2456 | 1124 | 5364 |
| mosGILT | Thiès | MONTH 12 | 2046 | 1236 | 3385 |
| mosGILT | Thiès | MONTH 18 | 1492 | 837 | 2659 |
| mosGILT | Thiès | MONTH 24 | 1790 | 974 | 3292 |
| mosGILT | Thiès | REINFECTION | 2743 | 268 | 28055 |
| mosGILT | Kédougou | DAY 0 | 289 | 136 | 613 |
| mosGILT | Kédougou | WEEK 2 | 391 | 204 | 749 |
| mosGILT | Kédougou | WEEK 4 | 315 | 163 | 610 |
| SAMSP1 | Thiès | DAY 0 | 1890 | 1307 | 2731 |
| SAMSP1 | Thiès | WEEK 2 | 2464 | 1826 | 3327 |
| SAMSP1 | Thiès | WEEK 4 | 2004 | 1289 | 3117 |
| SAMSP1 | Thiès | MONTH 3 | 1239 | 781 | 1965 |
| SAMSP1 | Thiès | MONTH 6 | 1943 | 1007 | 3750 |
| SAMSP1 | Thiès | MONTH 12 | 1485 | 945 | 2334 |
| SAMSP1 | Thiès | MONTH 18 | 1056 | 602 | 1853 |
| SAMSP1 | Thiès | MONTH 24 | 1314 | 726 | 2380 |
| SAMSP1 | Thiès | REINFECTION | 2120 | 140 | 32056 |
| SAMSP1 | Kédougou | DAY 0 | 449 | 204 | 989 |
| SAMSP1 | Kédougou | WEEK 2 | 539 | 250 | 1161 |
| SAMSP1 | Kédougou | WEEK 4 | 688 | 364 | 1300 |
